# Low SARS-CoV-2 infection rate and high vaccine-induced immunity among German healthcare workers at the end of the third wave of the COVID-19 pandemic

**DOI:** 10.1101/2021.08.02.21260667

**Authors:** Thomas Theo Brehm, Michelle Thompson, Felix Ullrich, Dorothee Schwinge, Marylyn M Addo, Anthea Spier, Johannes K Knobloch, Martin Aepfelbacher, Ansgar W Lohse, Marc Lütgehetmann, Julian Schulze zur Wiesch

**Affiliations:** I. Department of Internal Medicine, University Medical Center Hamburg-Eppendorf, Martinistraße 52, 20249 Hamburg, Germany; German Center for Infection Research (DZIF), Partner Site Hamburg-Lübeck-Borstel-Riems; Institute of Medical Microbiology, Virology and Hygiene, University Medical Center Hamburg-Eppendorf, Martinistraße 52, 20249 Hamburg, Germany

**Author notes:** contributed equally (Marc Lütgehetmann, Julian Schulze zur Wiesch). **Corresponding author:** Julian Schulze zur Wiesch, I. Department of Internal Medicine, University Medical Center Hamburg-Eppendorf, Martinistraße 52, 20246 Hamburg.

**Keywords:** SARS-CoV-2, COVID-19, healthcare workers, vaccination, immunity, Germany, seroprevalence

## Abstract

In this longitudinal cohort study, we assessed the severe acute respiratory syndrome coronavirus type 2 (SARS-CoV-2) seroconversion rates and analyzed the coronavirus disease 2019 (COVID-19) vaccine-induced immunity of 872 hospital workers at the University Medical Center Hamburg-Eppendorf between May 11 and May 31, 2021. The overall seroprevalence of anti-NC-SARS-CoV-2 antibodies was 4.7% (n=41), indicating low SARS-CoV-2 infection rates and persistent effectiveness of hospital-wide infection control interventions during the second and third wave of the pandemic. In total, 92.7% (n=808) out of the entire study cohort, 98.2% (n=325) of those who had been vaccinated once and all 393 individuals who had been vaccinated twice had detectable anti-S1-RBD-SARS-CoV-2 antibody titers and no significant differences in vaccine-induced immune response were detected between male and female individuals and between different age groups. Vaccinated study participants with detectable anti-NC-SARS-CoV-2 antibody titers (n=30) developed generally higher anti-S1-RBD-SARS-CoV-2 antibody titers compared to anti-NC-SARS-CoV-2 negative individuals (n=694) (median titer: 7812 vs. 345 BAU/ml, p<0.0001). Furthermore, study participants who received heterologous vaccination with AZD1222 followed by an mRNA vaccine showed markedly higher anti-S1-RBD-SARS-CoV-2 antibody titers than individuals who received two doses of an mRNA vaccine or two doses of AZD1222 (median titer: AZD1222 / AZD1222: 1069 BAU/ml, mRNA / mRNA: 1388 BAU/ml, AZD1222/mRNA: 9450 BAU/ml; p<0.0001). Our results demonstrate that infection control interventions were generally effective in preventing nosocomial transmission of SARS-CoV-2 and that COVID-19 vaccines can elicit strong humoral responses in the majority of a real-world cohort of hospital workers.

## Introduction

We have previously reported the first results of our severe acute respiratory syndrome coronavirus type 2 (SARS-CoV-2) seroprevalence project and could demonstrate low anti-S1-SARS-CoV-2 seroprevalence in 1253 hospital workers at the University Medical Center Hamburg-Eppendorf during the first wave of the coronavirus disease 2019 (COVID-19) epidemic up until July 2020 (Brehm et al., 2021c). As the epidemic in Germany evolved, an exponential increase in COVID-19 cases occurred during a second wave in autumn 2020 with incidence peaks with a 7-day incidence of more than 200 per 100,000 inhabitants in December and January 2020 and again during a third wave which peaked in April 2021 with more than 150 per 100,00 inhabitants (Robert-Koch-Institut, 2021a). At our center, the majority of COVID-19 patients were hospitalized during the second and third waves of the pandemic (Brehm et al., 2021a). Various infection control interventions such as universal masking, visitor restrictions, universal reverse transcription polymerase chain reaction (RT-PCR) admission screening of patients, and regular RT-PCR screening of asymptomatic healthcare workers (HCW) were rapidly implemented at our tertiary care center and repeatedly adapted throughout the epidemic. By June 2021, more than 150.000 SARS-CoV-2 RT-PCR tests were conducted among hospital employees, and a total of 111 infections were detected, the majority of which were classified as not work-related. However, it is not clear how many infections have been missed despite these screening efforts. Since both infection with SARS-CoV-2 and vaccination with the now licensed COVID-19 vaccines elicit anti-S1-RBD-SARS-CoV-2 antibodies, we adapted our strategy to detect resolved infections by screening for the anti-NC-SARS-CoV-2 response. By June 2021, the European Medicines Agency (EMA) has granted conditional marketing authorizations for the two mRNA COVID-19 vaccines BNT162b2 (Comirnaty, Biontech/Pfizer) (EMA, 2021a), mRNA-1273 (Moderna / NIAID) (EMA, 2021b), and the viral vector-based vaccines AZD1222 (Vaxzevria, AstraZeneca) (EMA, 2021c), and Ad26.COV2.S (Janssen) (EMA, 2021d). The respective phase 3 trials reported high efficacy in priming neutralizing anti-Spike-SARS-CoV-2 antibodies and preventing symptomatic SARS-CoV-2 infections after a single dose (AZD1222) (Sadoff et al., 2021) or two doses administered three (BNT162b2) (Polack et al., 2020), four (mRNA-1273) (Baden et al., 2021) or 12 weeks (AZD1222) (Voysey et al., 2021) apart. However, vaccine efficacy may differ in different populations, and vaccine regimens may vary depending on availability, national guidelines, and findings of post-marketing surveillance. Since the administration of AZD1222 was suspended in individuals below 60 years in Germany after the occurrence of vaccine-induced immune thrombotic thrombocytopenia in March 2021 (Greinacher et al., 2021; Schultz et al., 2021), heterologous boosting with an mRNA vaccine was recommended in this group of vaccinees. As evidence on the short and mid-term immune responses to these different COVID-19 vaccine regimens is currently limited, real-world studies are urgently needed to develop rational and efficient vaccination schedules for the long-term protection of both hospital employees and their patients. As part of our seroprevalence project, we performed another study visit to determine the number of hospital employees who were knowingly or unknowingly infected with SARS-CoV-2 as well as to assess the vaccine-induced humoral immunity to different COVID-19 vaccine regimens.

## Materials and methods

### Study design

Participants of our ongoing SARS-CoV-2 seroprevalence study were recruited by informing employees of the University Medical Center Hamburg-Eppendorf and written informed consent was obtained by all study participants before recruitment. For the present study visit, participants were invited to provide a serum sample between May 11 and May 31, 2021. Demographic and occupational characteristics, prior SARS-CoV-2 infections, and vaccination status were assessed using an online REDcap electronic data capture tool specifically designed for the present study. The two licensed mRNA vaccines, BNT162b2 and mRNA-1273, were subsequently subsumed to one category. The study protocol was reviewed and approved by the Ethics Committee of the Medical Council of Hamburg (PV 7298).

### Serology

To differentiate between past infection and vaccine-induced immune response, anti-SARS-CoV-2 antibodies against the viral nucleocapsid (NC) and the receptor-binding domain (RBD) domain of the viral spike protein (S) were determined using the qualitative anti-NC-SARS-CoV-2 Ig assay (Elecsys Anti-SARS-CoV-2, Roche, Mannheim Germany; cut off ≥ 1 COI/ml) and the quantitative anti-S1-RBD-SARS-CoV-2 assay (Elecsys Anti-SARS-CoV-2 Spike, Roche, Mannheim, Germany; cut off 0.8 U/ml). Both electrochemiluminescence immunoassays (Cobas e411, Roche; Mannheim, Germany) use a double-antigen sandwich assay format that detects IgA, IgM, and IgG. Samples with titers above 250 U/ml were manually diluted 1:100 in dilution buffer according to the manufacturer’s recommendations to increase the linear range to 25000 U/ml. A mathematical transposition of Elecsys anti-S1-RBD-SARS-CoV-2 specific U/ml to the World Health Organization (WHO) standard BAU/ml was performed using the following equation: U/ml□=□0.972 x BAU/ml (Resman Rus et al., 2021). Since it has been previously demonstrated that after natural infection, an anti-S1-RBD-SARS-CoV-2 antibody concentration of more than 133 BAU/ml predicts the presence of neutralizing antibodies (Resman Rus et al., 2021) and may thus be a potential surrogate for high protection from COVID-19 after vaccination, we specifically calculated the percentage of different subgroups of our study cohort with antibody titers above this threshold.

### Statistical analyses

A two-tailed Mann–Whitney U test was used to analyze the median antibody titers between subgroups. In addition, linear regression analysis was performed to assess the correlation between age and antibody titers. P values less than 0.05 are considered statistically significant. Statistical analyses were performed using GraphPad Prism, version 9 for macOS (GraphPad Software, La Jolla, California, USA).

## Results

### Characterization of the study population

A total of 872 individuals participated in the current study visit, representing around 8% of all employees of the University Medical Center Hamburg-Eppendorf. The median age was 38 years (IQR 30 – 49 years), and 78.0% (n=680) of the study population were women (**Table 1**). The majority of study participants were HCW (81.7%, n=712) directly involved in patient care at different departments of our tertiary care center: 36.0% were nurses (n=314), 20.1% physicians (n=175), 10.1% medical technicians (n=88) and 15.5% had other professions (n=135). Information on prior SARS-CoV-2 infections and vaccination status was provided by the majority of study participants (90.6%, n=790) (**Supplemental Table 1**). Of those, 41.9% (n=331) had received one dose of a COVID-19 vaccine (AZD1222: n=267, mRNA: n=64). Another 49.7% (n=393) had received two COVID-19 vaccine doses (AZD1222 / AZD1222: n=25; AZD1222 / mRNA: n=106; mRNA / mRNA: n=261) at the time of the study visit. Thirty-one individuals (3.9%) reported that they had been diagnosed with SARS-CoV-2 in the past.

**Table 1.**
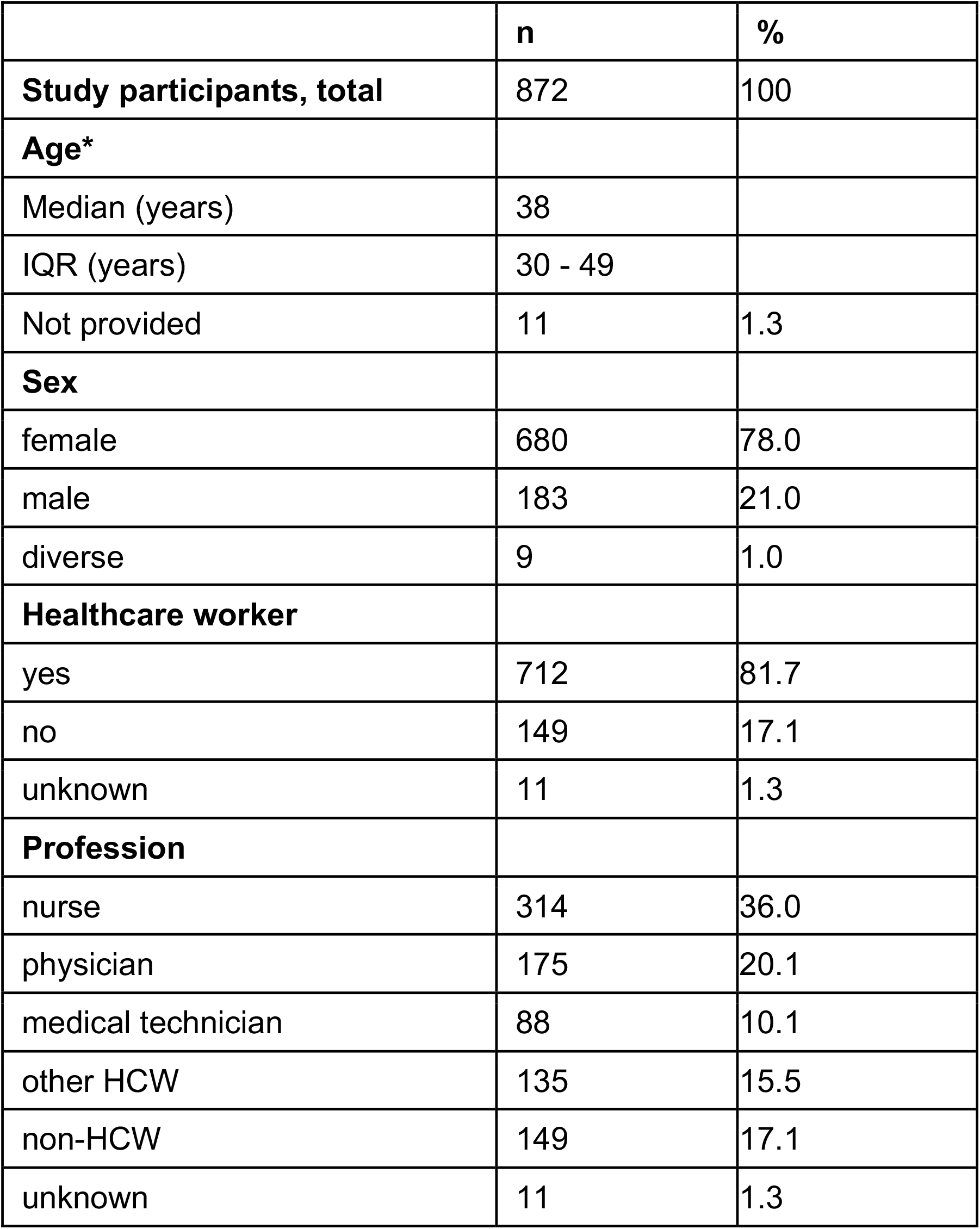
Characterization of the study cohort

|  | n | % |
| --- | --- | --- |
| **Study participants, total** | 872 | 100 |
| **Age\*** |  |  |
| Median (years) | 38 |  |
| IQR (years) | 30 - 49 |  |
| Not provided | 11 | 1.3 |
| **Sex** |  |  |
| female | 680 | 78.0 |
| male | 183 | 21.0 |
| diverse | 9 | 1.0 |
| **Healthcare worker** |  |  |
| yes | 712 | 81.7 |
| no | 149 | 17.1 |
| unknown | 11 | 1.3 |
| **Profession** |  |  |
| nurse | 314 | 36.0 |
| physician | 175 | 20.1 |
| medical technician | 88 | 10.1 |
| other HCW | 135 | 15.5 |
| non-HCW | 149 | 17.1 |
| unknown | 11 | 1.3 |

### Serological results

At the current study visit, 4.7% (n=41) of the study population had detectable anti-NP-antibodies indicating prior infection with SARS-CoV-2. Of those, 31 (76%) had been knowingly infected with SARS-CoV-2 in the past, while 12 (29%) did not report a previous infection. In total, 92.7% (n=808) out of the entire study cohort, 98.2% (n=325) of those who had been vaccinated once and all 393 individuals who had been vaccinated twice had detectable anti-S1-RBD-SARS-CoV-2 antibody titers. Among all vaccinated study participants, those with detectable anti-NC-SARS-CoV-2 antibody titers (n=30) developed generally higher anti-S1-RBD-SARS-CoV-2 antibody titers compared to anti-NC-SARS-CoV-2 negative individuals (n=694) (median titer: 7812 vs. 345 BAU/ml, p<0.0001). Among those individuals who had received one vaccine dose with AZD1222, those who were anti-NC-SARS-CoV-2 positive (median titer: 7797 BAU/ml) had significantly higher anti-S1-RBD-SARS-CoV-2 antibody titers than those who were anti-NC-SARS-CoV-2 negative (median titer: 61 BAU/ml; p<0,0001) (**Figure 1**). Likewise, anti-S1-RBD-SARS-CoV-2 antibody titers were higher amongst individuals with detectable anti-NC-SARS-CoV-2 antibodies compared to those without detectable anti-NC-SARS-CoV-2 antibodies both in the subgroup of individuals who had received one dose of an mRNA vaccine (median titer: 15269 BAU/ml vs 109 BAU/ml; p<0.0001) and the subgroup that had received two doses of an mRNA vaccine (median titer; 4725 BAU/ml vs 1333 BAU/ml; p=0.0003). In subgroups with other vaccination regimens, the number of individuals with detectable anti-NC-SARS-CoV-2 antibodies was too small for detailed statistical analysis. Since the interval between the first and the second vaccine at our center was generally six weeks for mRNA vaccines and 12 weeks for AZD1222, the duration after the first vaccine dose at the time of the study visit was generally longer amongst individuals who had received one dose of AZD1222 (median duration: AZD1222: 80 days, mRNA: 28 days, p<0.0001), but anti-S1-RBD-SARS-CoV-2 antibody titers were higher among individuals who had received an mRNA vaccine (median titer: AZD1222: 63 BAU/ml, mRNA: 122 BAU/ml; p=0.005) (**Figure 2A and 2B**). Among study participants who had already received two vaccine doses, the duration after the second dose was markedly higher amongst those who had received two doses of an mRNA vaccine since those vaccines were licensed and available earlier compared to AZD1222 (median duration: AZD1222 / AZD1222: 9 days, mRNA / mRNA: 106 days, AZD1222 / mRNA: 9 days; p<0.001) (**Figure 2 C and 2D**). Of note, the study participants who received vaccination with AZD1222 followed by an mRNA vaccine showed significantly higher anti-S1-RBD-SARS-CoV-2 antibody titers than individuals who had received two doses of an mRNA vaccine or two doses of AZD1222 (median titer: AZD1222 / AZD1222: 1069 BAU/ml, mRNA / mRNA: 1388 BAU/ml, AZD1222 / mRNA: 9450 BAU/ml; p<0.0001). Anti-S1-RBD-SARS-CoV-2 antibody concentrations above the cut-off of 133 BAU/ml, which has been shown to predict the presence of neutralizing antibodies after natural SARS-CoV-2 infections (Resman Rus et al., 2021), were detected in 28.7% (n=95) of those who received one vaccine dose and 93.6% (n=368) who received two vaccine doses. COVID-19 vaccinees are generally regarded fully vaccinated two weeks after the second vaccination (Keehner et al., 2021), at which time all but two study participants (99.3%, n=287) had antibody titers above 133 BAU/ml (**Supplemental Figure 1**). Linear regression analysis did not demonstrate a significant correlation between age and anti-S1-RBD-SARS-CoV-2 titers amongst study participants who had received one (R^2^=0.02) or two (R^2^=0.001) vaccine doses (**Supplemental Figure 2**). Median anti-S1-RBD-SARS-CoV-2 antibody titers did not significantly differ between male and female study participants after the first (median titer: 52 BAU/ml vs. 74 BAU/ml; p=0.07) and the second (1456 BAU/ml vs. 1705 BAU/ml; P=0.98) vaccine dose.

**Figure 1.**
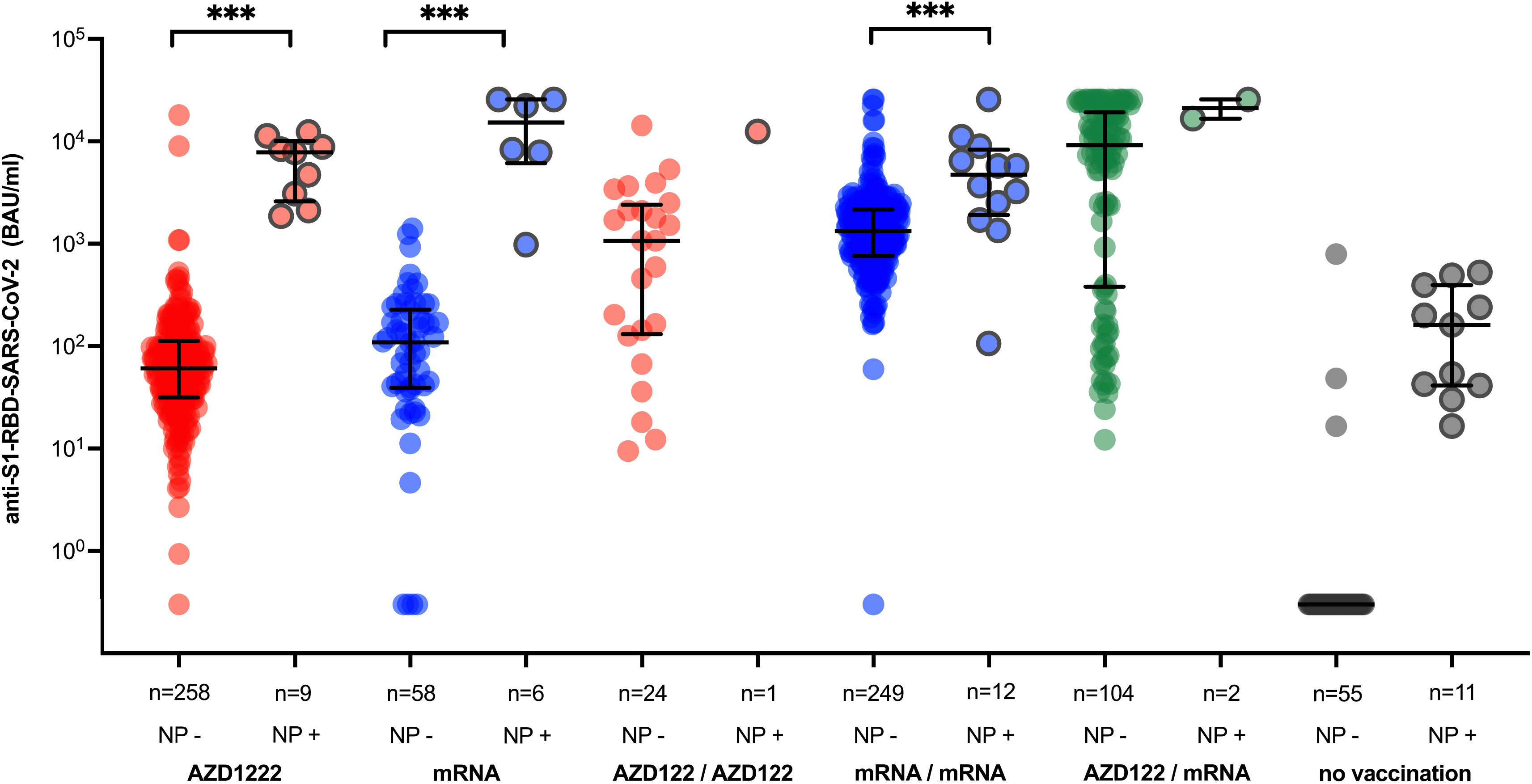
Anti-S1-RBD-SARS-CoV-2 antibody titers based on infection status and different vaccination regimens. Anti-S1-RBD-SARS-CoV-2 antibody titers on a logarithmic scale among study participants with (NP+) and without (NP-) detectable anti-NC-SARS-CoV-2 based on their vaccination status. Individual data points are shown as an aligned dot plot with lines showing the median with the interquartile range. Significant differences were determined by two-tailed Mann–Whitney U test (***P□<□0.001; **P□<□0.01; *P□<□0.05).

**Figure 2.**
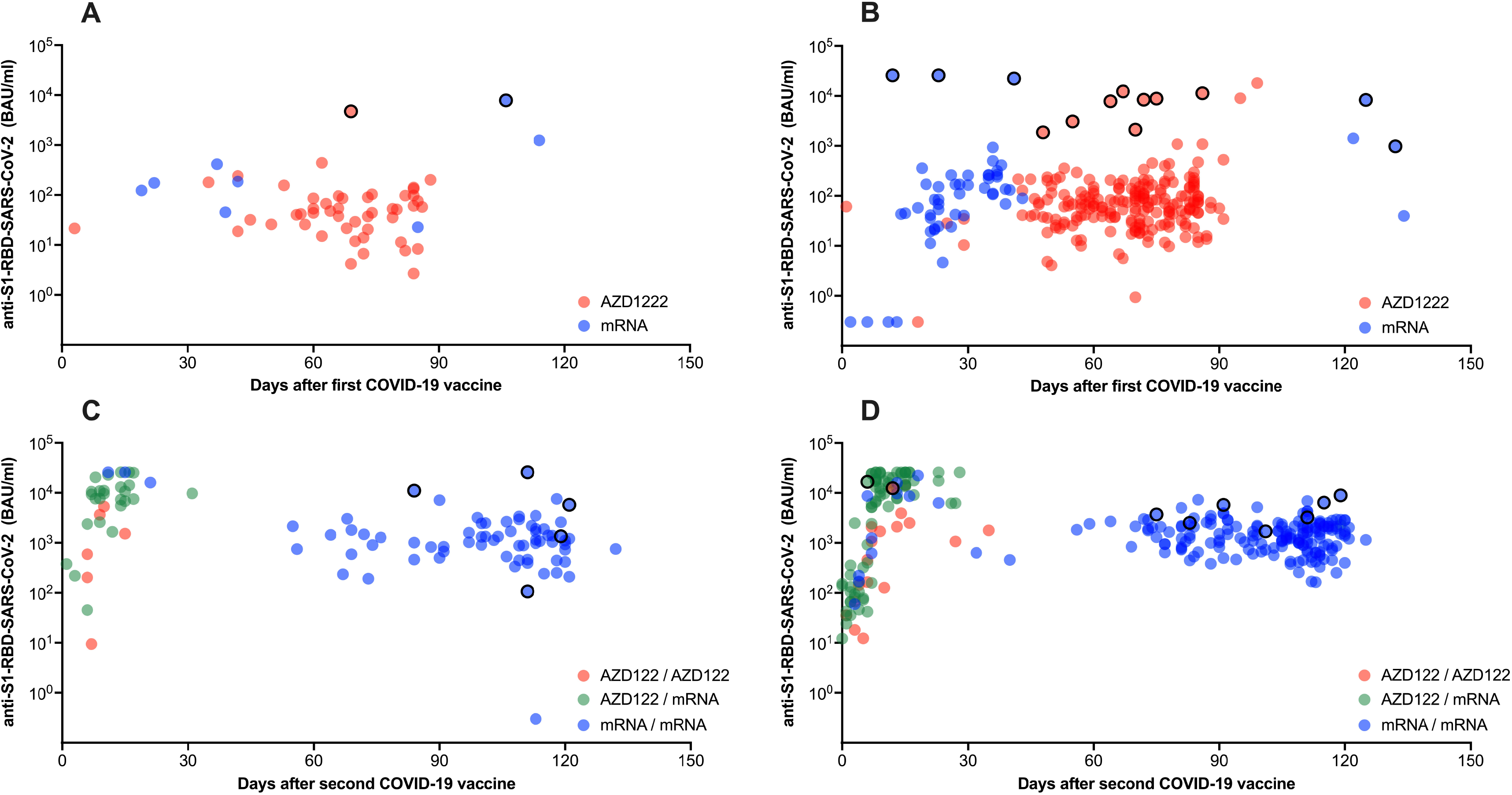
Anti-S1-RBD-SARS-CoV-2 antibody titers based on sex, vaccination status and duration after vaccination. Anti-S1-RBD-SARS-CoV-2 antibody titers on a logarithmic scale among male (A, C) and female (B, D) study participants with (dots with black edging) and without (dots without black edging) detectable anti-NC-SARS-CoV-2 plotted against the duration after administration after the first (A, B) and second (B, C) COVID-19 vaccine dose.

## Discussion

Here we describe the prevalence and titer of anti-S1-RBD-SARS-CoV-2 and anti-NC-SARS-CoV-2 antibodies in 872 hospital workers at our German tertiary care center in May 2021 based on their vaccination and infection status.

We have previously reported an anti-S1--SARS-CoV-2 seroprevalence of 1.8% among a cohort of hospital workers at our center by the end of the first wave in early July 2020 (Brehm et al., 2021c). One of the main findings of the current study visit was that only 4.7% of the hospital employees had detectable anti-NC-SARS-CoV-2 antibodies, indicating persistently low nosocomial SARS-CoV-2 transmission throughout the second and third wave of the COVID-19 epidemic. Since the official rate of recovered COVID-19 patients among inhabitants of the city-state of Hamburg was 4.2% in June 2021 and further unidentified cases have to be assumed, the infection rate among hospital employees appears to be not increased compared to the general population. The fact that only a few infections in hospital workers occurred during the second and third wave of the COVID-19 pandemic despite a large number of patients treated at our center (Brehm et al., 2021a) indicate the persistent effectiveness of the progressive infection control interventions that included universal RT-PCR admission screening of patients and regular RT-PCR screening of HCW. These findings are reassuring since, despite increasing vaccination coverage, both hospital workers and vulnerable patient groups will have to be protected from nosocomial SARS-CoV-2 transmission in the upcoming months.

Another important finding of our study is the fact that the vast majority of our study cohort developed anti-S1-RBD-SARS-CoV-2 antibody titers that have been shown to correlate with neutralizing activity (Resman Rus et al., 2021) after the second vaccine dose irrespective of age, sex, and respective vaccine regimen. While these observations are reassuring, a reliable absolute antibody threshold for individual protection from COVID-19 may not exist, and infections may occur even in the presence of neutralizing antibodies (Brehm et al., 2021b). However, population-based correlates for immunity appear attainable and need to be identified by large prospective studies in the future (Jin et al., 2021). All study participants obtained their individual serological results as well as the results of the entire study cohort and were given a chance to discuss these findings with the scientific study team. We feel that these interactions provided reassurance to the hospital employees about the generally high potency of COVID-19 vaccines on the one hand and the limited predictive value of individual antibody titers for the level of protection from SARS-CoV-2 on the other hand.

Strikingly, study participants receiving a heterologous prime-boost vaccination regimen with AZD1222 vaccine followed by an mRNA vaccine developed significantly higher antibody titers than individuals receiving homologous vaccination regimens. This finding adds to preliminary data indicating that heterologous vaccination regimens may be more immunogenic than homologous vaccine regimens and have comparable reactogenicity profiles (Hillus et al., 2021; Schmidt et al., 2021; Borobia et al., 2021). While these results still need further confirmation in larger cohorts, these findings may have implications for future vaccine strategies against COVID-19 and other pathogens.

It has been previously demonstrated that the humoral immune response to a single COVID-19 vaccine dose in individuals primed by natural infection may provide antibody titers at least comparable to those found in non-infected individuals after the second vaccine dose (Blain et al., 2021; Havervall et al., 2021; Krammer et al., 2021; Reynolds et al., 2021; Saadat et al., 2021). In our study cohort, unvaccinated participants with detectable anti-NC-SARS-CoV-2 antibodies indicating prior infection had lower anti-S1-RBD-SARS-CoV-2 antibody compared to anti-NC-SARS-CoV-2 negative individuals who were vaccinated twice, indicating that booster vaccination is indeed needed for those recovered patients. However, anti-NC-SARS-CoV-2 positive individuals developed markedly higher anti-S1-RBD-SARS-CoV-2 antibody titers after both one and two vaccine doses compared to anti-NC-SARS-CoV-2 negative individuals regardless of the vaccine regimen. While Germany’s official public health institute currently recommends the administration of only a single dose of any of the COVID-19 vaccines for individuals with prior SARS-CoV-2 infection (Robert-Koch-Institut, 2021b), the duration of protection as well as the optimal timing of booster doses and selection of the most effective vaccine regimen yet need to be determined.

Our study has important limitations, most of which are inherent to the cross-sectional character of our study. First, we only provide vaccine-induced antibody titers for point in time and are not able to assess longitudinal data or serological kinetics. Also, since different COVID-19 vaccines were licensed and distributed to hospital workers at different points in time, we are not able to compare immune responses elicited by different vaccines at the same post-vaccination interval. Second, while our longitudinal study was initiated before COVID-19 vaccines could be anticipated, and study participants were recruited across all age groups and occupations, we did not recruit a strictly representative sample of hospital employees at our tertiary care center, which limits the overall generalizability of results. Study participants with a positive attitude towards COVID-19 vaccines in general and those who have been vaccinated in particular may be more likely to participate in our study which may represent an important selection bias. Third, not all individuals develop a long-lasting anti-NC-SARS-CoV-2 response follow infection with SARS-CoV-2 (Van Elslande et al., 2021), so we may underestimate the true overall infection rate in our cohort. Fourth, the number of study participants, especially those with anti-NC-SARS-CoV-2 antibodies, was relatively small in some subgroups, and our findings need to be confirmed by larger cohort studies.

While the COVID-19 pandemic evolves and the overall vaccination coverage increases, prospective longitudinal studies will be necessary to get a better understanding of the kinetics humoral and cellular immune responses to different COVID-19 vaccines, to identify correlates of protection against SARS-CoV-2, and to determine the duration and quality of infection- and vaccine-induced immunity. In light of the increasing occurrence of variants of concern, it will be important to determine how different vaccination regimens may influence protection against COVID-19.

## Conclusion

At our tertiary care center, the number of SARS-CoV-2 infections among exposed hospital employees remained low throughout the second and third waves of the pandemic. Anti-S1-RBD-SARS-CoV-2 antibody titers show a high rate of strong humoral response to COVID-19 vaccines regardless of age, sex, and vaccine regimens and indicate a high level of protection from SARS-CoV-2 infections in our real-world cohort. Significantly higher anti-S1-RBD-SARS-CoV-2 antibody titers among individuals receiving heterologous prime-boost vaccination regimens with AZD1222 followed by an mRNA vaccine may suggest even higher efficacy than homologous regimens.

## Data Availability

N/A

## Acknowledgments

We thank Nils Dittberner, Sabrina Kreß and Martina Fahl for excellent technical assistance. We thank all study participants and departments of the University Medical Center Hamburg-Eppendorf for their active participation in the study.

## Figure titles and legends

**Supplemental Figure 1**

**Title:** Anti-S1-RBD-SARS-CoV-2 antibody titers of study participants more than 14 days after the second vaccine dose

**Legend:** Anti-S1-RBD-SARS-CoV-2 antibody titers on a logarithmic scale among study participants who received their second vaccine dose at least 14 days before the study visit with (NP+) and without (NP-) detectable anti-NC-SARS-CoV-2 based on the vaccination regimen. Individual data points are shown as an aligned dot plot with lines showing the median with the interquartile range.

**Supplemental Figure 2**

**Title:** Anti-S1-RBD-SARS-CoV-2 antibody titers and age

**Legend:** Anti-S1-RBD-SARS-CoV-2 antibody titers on a logarithmic scale after the first (A) and second (B) vaccine dose plotted against the age of study participants. Linear regression analysis was performed to assess the correlation between age and antibody titers.

